# Spatiotemporal tools for emerging and endemic disease hotspots in small areas – an analysis of dengue and chikungunya in Barbados, 2013 – 2016

**DOI:** 10.1101/19011262

**Authors:** Catherine A. Lippi, Anna M. Stewart-Ibarra, Moory Romero, Avery Q.J. Hinds, Rachel Lowe, Roché Mahon, Cedric J. Van Meerbeeck, Leslie Rollock, Marquita Gittens-St. Hilaire, Adrian R. Trotman, Dale Holligan, Shane Kirton, Mercy J. Borbor-Cordova, Sadie J. Ryan

## Abstract

**Objective:** To detect potential hotspots of transmission of dengue and chikungunya in Barbados, and assess impact of input surveillance data and methodology on observed patterns of risk.

**Methods:** Using two methods of cluster detection, Moran’s I and spatial scan statistics, we analyzed the geospatial and temporal distribution of disease cases and rates across Barbados for dengue fever in 2013–2016, and a 2014 chikungunya outbreak.

**Results:** During years with high numbers of dengue cases, hotspots for cases were found with Moran’s I in south and central regions in 2013 and 2016, respectively. Using smoothed disease rates, clustering was detected every year for dengue. Hotspots were not detected via spatial scan statistics, but coldspots suggesting lower rates of disease activity were found in southwestern Barbados during high case years of dengue.

**Conclusions:** Spatial analysis of surveillance data is useful in identifying outbreak hotspots, complementing existing early warning systems. We caution that these methods should be used in a manner appropriate to available data, and reflecting explicit public health goals – managing for overall case numbers, or targeting anomalous rates for further investigation.

## Introduction

Dengue fever threatens the health of communities throughout Latin America and the Caribbean, where all four serotypes of dengue virus (DENV 1–4) are in circulation following a regional resurgence of the pathogen in the 1980s ^1,2^. The Caribbean island of Barbados is challenged with managing endemic dengue fever, and other febrile mosquito-borne diseases including emerging chikungunya and Zika viral diseases ^3,4^. In small island nations like Barbados, outbreaks translate into increased morbidity and mortality, high costs to healthcare systems, and lost economic productivity ^5–7^. With approximately 40% of employment and gross domestic product linked to the tourism industry, Barbados is particularly vulnerable to the economic impacts of arbovirus outbreaks ^8^. In addition to lost domestic productivity, travel-related cases and negative health perceptions associated with outbreaks deter potential visitors, further impacting the livelihoods of island residents ^9,10^. The emergence and subsequent establishment of novel arboviruses in the Caribbean exacerbates matters by complicating disease management while further impacting sources of income ^3^. In response to these social and economic burdens, the Ministry of Health and Wellness of Barbados (MoH) has a long history of engaging in public mosquito control and active disease surveillance, where suspected human cases are laboratory confirmed, and vector control interventions are conducted in response to both lab results and mosquito surveillance. Interagency collaborations are part of a comprehensive effort to mitigate the toll of endemic dengue^11^. Previous studies performed in Barbados have described climatological and seasonal drivers of dengue transmission, vital components of early warning systems and forecasting models ^12^. While large-scale climatological factors undoubtedly play a dominant role in driving outbreaks of mosquito-borne illness, this plays out at the local scale as a function of the human landscape ^13,14^. Therefore understanding the local distribution of human cases is also necessary for understanding patterns of exposure risk and guiding vector abatement strategies.

*Aedes aegypti* is the primary mosquito vector of medical concern throughout the Caribbean. Public health vector control programs are widely acknowledged as cost effective in controlling arboviruses transmitted by *Ae. aegypti*, relative to costs associated with the delivery of health services and supportive care ^15^. Nevertheless, public health resources are finite, calling for efficient intervention strategies to target mosquito populations and suppress transmission pathways. *Aedes aegypti* is a container-breeding mosquito, and successfully exploits anthropogenic environments for oviposition and larval rearing. The role of household-level characteristics, such as housing condition and water storage habits, in promoting mosquito production has been repeatedly demonstrated^13,14^. In some instances, favorable microhabitats enable mosquitoes, and subsequently disease transmission, to persist in spite of generally unfavorable environmental conditions ^16^. Thus, identifying spatial clusters of high disease activity, or “hotspots,” can prove invaluable when prioritizing the delivery of abatement and outreach services. Further additional challenges, while not unique in the context of integrated vector control, are essential to address for management of mosquito-borne diseases in Caribbean islands. Vector-borne disease risk can shift rapidly on small island like Barbados due to many factors including insecticide resistance, climate variability, climate change, high disease prevalence, and variable mosquito control efforts in response to herd immunity dynamics. Small island developing states in the Caribbean also face challenges to the elimination of *Ae. aegypti*, as reintroductions of pathogens and vectors are frequent due to inter-regional travel, unplanned urbanization, and limited resources for vector control ^17^. These management challenges demand strategies that incorporate spatially and temporally sensitive methods of detecting transmission activity.

Geographic information systems (GIS) offer a powerful tool in the visualization and incorporation of spatial epidemiological data into public health programs ^18^. While many health departments and ministries have readily adopted GIS methods into their surveillance and reporting activities, fewer have extended these methodologies to incorporate statistical tests of spatial dependency in human case data. Local Indicators of Spatial Association (LISA) statistics are routinely used in an exploratory framework to quantitatively describe patterns of spatial dependence and clustering, or dispersion, of disease cases within a defined area of study ^19^. Identifying spatially discrete areas of significantly high (i.e. hotspots), or low (i.e. coldspots), disease activity within functional administrative boundaries is a useful framework for crafting responses to outbreak events and future interventions, enabling agencies to focus their efforts more efficiently. Global and local Moran’s I tests have been applied in public health contexts to describe spatial distributions of mosquito-borne disease outbreaks, including dengue fever, and to detect the location of disease clusters ^14,20,21^.

While LISA methods give us insight into the spatial structure of disease activity within a given time period, these analyses are temporally static. In instances where georeferenced disease surveillance data are available at regular time intervals, spatial scan statistics can be employed to identify local areas of clustering in multiple dimensions (i.e. space, time, or space-time). Spatial scan statistics are capable of detecting possible disease clustering in case-only surveillance data, using a series of variable search windows to evaluate spatial and temporal trends in the dataset ^22,23^. Application of space-time scan statistics can be a powerful tool in disease surveillance and outbreak detection, where we are interested in describing not only where, but also when clusters of events occur over a continuous period of time.

To our knowledge, no previous efforts have described the spatial and temporal distribution of dengue or chikungunya outbreaks in Barbados. Using epidemiological case data collected by the MoH in Barbados from 2013 – 2016, we used exploratory LISA and space-time scan statistics to test for spatial and temporal autocorrelation of dengue and chikungunya cases within operational health districts. The objectives of this study were to i) detect global spatial dependency, or clustering, of surveillance arbovirus cases reported in Barbados within each year of the study period; ii) when global spatial dependency is detected, identify the locations of hotspots (i.e. clustered) and coldspots (i.e. dispersed) of disease activity; iii) assess the effect of different input data on observed spatial patterns; and iv) detect spatiotemporal patterns in disease rates at finer temporal resolutions via spatial scan statistics. This also provides an important opportunity to discuss and showcase the implications of how these methods are implemented in situations where data are limited, simply as a function of small populations, as seen in small island nations.

## Methods

### Study Area and Epidemiological Data

Barbados, situated in the Caribbean, has an estimated residential population of over 277,000 ^24^. The most densely populated areas are found on the southern side of the island, with the highest population density found around Bridgetown, the capital city ^24^. Transmission of mosquito-borne diseases in Barbados is seasonal, with peak transmission typically associated with high numbers of mosquitoes during the rainy season (June –November), and fewer disease cases reported during the dry season (December–May) ^12,25^. The MoH of Barbados performs active and passive surveillance for dengue and other mosquito-borne diseases via nine polyclinics. These serve seven polyclinic administrative catchment (PAC) areas (Branford Taitt, David Thompson, Eunice Gibson, Maurice Byer, Randall Philip, St. Philip, and Winston Scott), which are further divided into 63 health districts (Fig. 1).

**Fig. 1.**
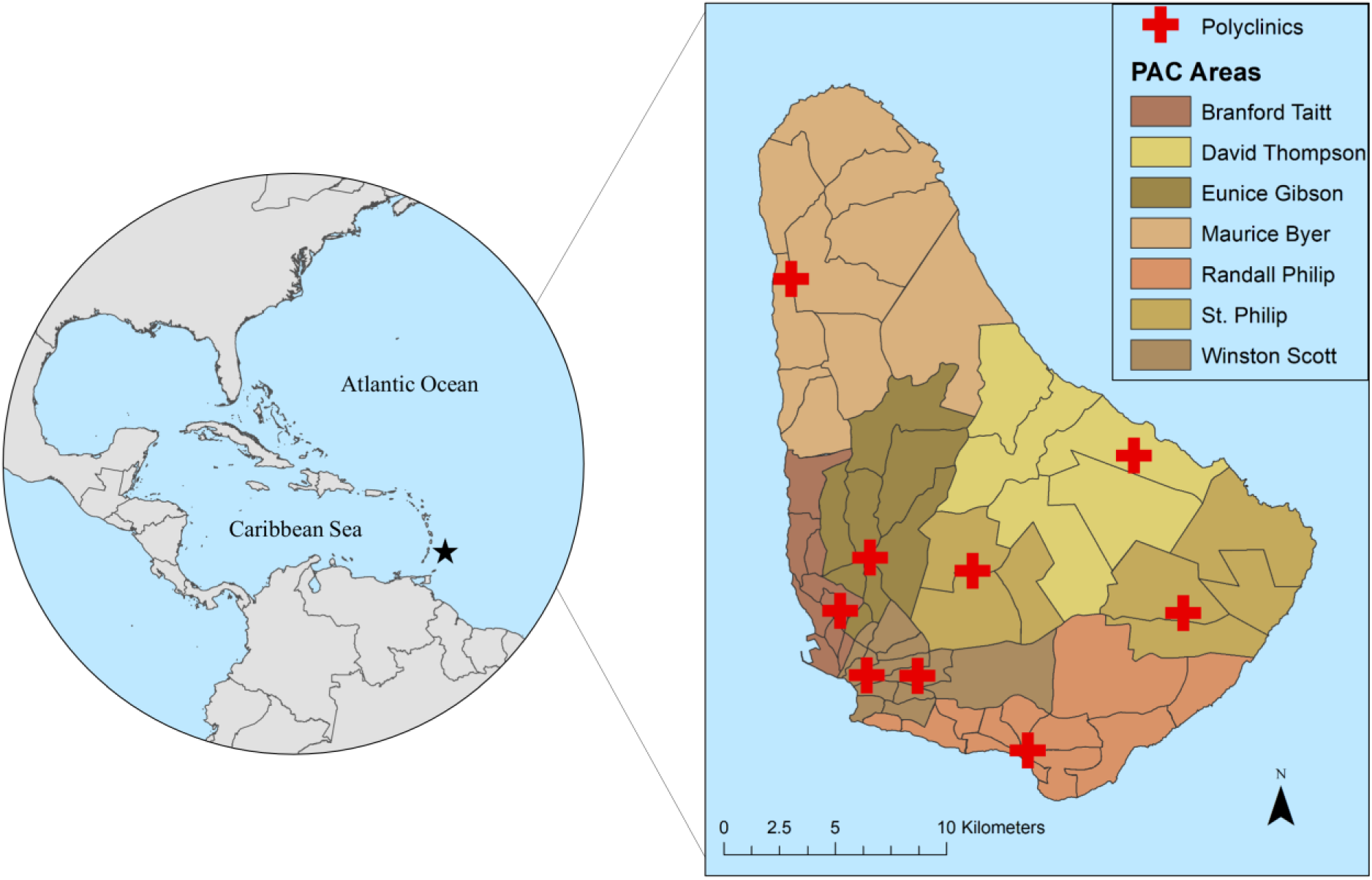
Health districts and polyclinic administrative catchment (PAC) areas in Barbados. This figure was produced in ArcMap 10.4 (ESRI, Redlands, CA) using shapefiles from the GADM database of Global Administrative Areas, ver. 2.8 (gadm.org), and shapefiles provided by the MoH, Barbados.

Public vector control and health services are delivered at the level of health districts, which range in size from 0.40 km^2^ to 26.62 km^2^. The MoH oversees arbovirus sureveillance activities, where suspected human cases of dengue and chikungunya are recorded by the Ministry and confirmed in the National Reference Laboratory by RT-PCR or ELISA. De-identified, monthly case totals for dengue fever in each health district were provided by the MoH for the years 2013 – 2016, and we defined the location of cases as the centroid of a given district. Georeferenced data on lab-confirmed chikungunya cases, aggregated to health districts, were also made available for this study, but were only available for the 2014 outbreak. Additional GIS data were provided for this study by the MoH, including shapefiles of the administrative boundaries for health districts in Barbados.

### Global and Local Indicators of Spatial Association

Annual case totals for dengue and chikungunya in each health district were aggregated from monthly case data provided by the MoH for each year of the study. Annual per capita disease rates were derived from annual totals and population data from the most recent national census, conducted in 2010 ^24^. The population of each health district ranged from 68 to 12,743, according to census data. Due to low population in some health districts, raw disease rates may be susceptible to instability due to high variance associated with small numerators or denominators (i.e. the “small number problem”) ^26^. Performing spatial analyses on raw rates with high instability can result in incorrectly identifying artefacts of the small number problem as statistically significant outliers. We performed Empirical Bayes smoothing (EB), where the variance of rate estimates is globally reduced via *a priori* probability functions, on raw disease rates in Geoda (ver. 1.12.0) to compensate for high variability in rates due to low health district population. EB smoothed rates were compared to raw disease rates, verifying the overall reduction of variance from smoothing.

Global Moran’s I with inverse distance weighting (ArcMap, ver. 10.4) was used to test for spatial autocorrelation in both case counts and smoothed disease rates for dengue and chikungunya in Barbados for each year of the study. A global indicator of spatial dependence, the Moran’s I statistic provides a measure of the degree of statistically significant clustering or dispersion in disease measures for the entire island. Local Moran’s I is a LISA statistic for identifying locations (e.g. health districts within the study area) with statistically similar spatial patterns (e.g. clustering or dispersion) of high and low values (i.e. hotspots or coldspots) on the island ^27^. This statistic is also useful for the detection of spatial outliers, locations with significantly extreme values compared to neighboring areas ^27^. Local Moran’s I with inverse distance weighting was performed for each reported year in ArcMap (ver. 10.4) to identify health districts that were hotspots, or coldspots, of dengue or chikungunya activity.

### Spatial Scan Statistics

We compared the spatial distribution of dengue and chikungunya found via LISA statistical analyses, calculated for each year of the study, to patterns of clustering and dispersion in continuous aggregated cases over the study period. Patterns of spatiotemporal clustering in monthly case totals within each year were tested using the univariate Kuldorrff space-time scan statistics in SaTScan (ver. 9.4.4), where we performed retrospective space-time analyses, scanning for both clustering and dispersion ^28^. A circular search window was specified to test for spatiotemporal clustering, comparing cases at each location (i.e. centroids of health districts) to neighboring areas within a variable window, using a time precision of one month. Clusters were constrained to a maximum cluster size of 50% of case data, a maximum temporal window of 50% of the study period, and allowed for geographic overlap with other clusters, provided that no neighboring cluster centers were included in a given cluster. Likelihood ratios and associated p-values were reported for each identified cluster, where maximum likelihood values were calculated via Monte Carlo simulation (999 replications). Statistically significant clusters (α=0.05) from the SaTScan analyses were mapped with LISA results for each year in ArcMap (ver. 10.4) for visual comparison.

## Results

The number of dengue cases in Barbados reported by the MoH fluctuated greatly during the study period, with large outbreaks occurring in 2013 (n=526) and 2016 (n=386), and lower case numbers in 2014 (n=147) and 2015 (n=58). Georeferenced cases of chikungunya (n=57) were only available for 2014. We detected statistically significant (α=0.05) global clustering (i.e. Moran’s I values > 0) in aggregated case counts during the years of large dengue outbreaks, 2013 and 2016 (Table 1), while significant global clustering of EB smoothed rates was found in every year for dengue (Table 2). No significant clustering was detected during the 2014 chikungunya outbreak.

**Table 1.** Global Moran’s I values for dengue and chikungunya case totals, aggregated to health district, in each year in the study.

| Arbovirus | Year | Total Cases | Moran's I<sup>†</sup> | Z-Score | P-value |
| --- | --- | --- | --- | --- | --- |
| Dengue | 2013 | 526 | 0.23 | 4.40 | **< 0.001\*** |
| Dengue | 2014 | 147 | -0.04 | -0.51 | 0.610 |
| Dengue | 2015 | 58 | 0.04 | 1.06 | 0.291 |
| Dengue | 2016 | 386 | 0.11 | 2.29 | **0.022\*** |
| Chikungunya | 2014 | 57 | 0.06 | 0.84 | 0.400 |

<sup>†</sup> Moran's I values range between -1 and 1, where negative values indicate dispersion and positive values indicate clustering.

**Table 2.**
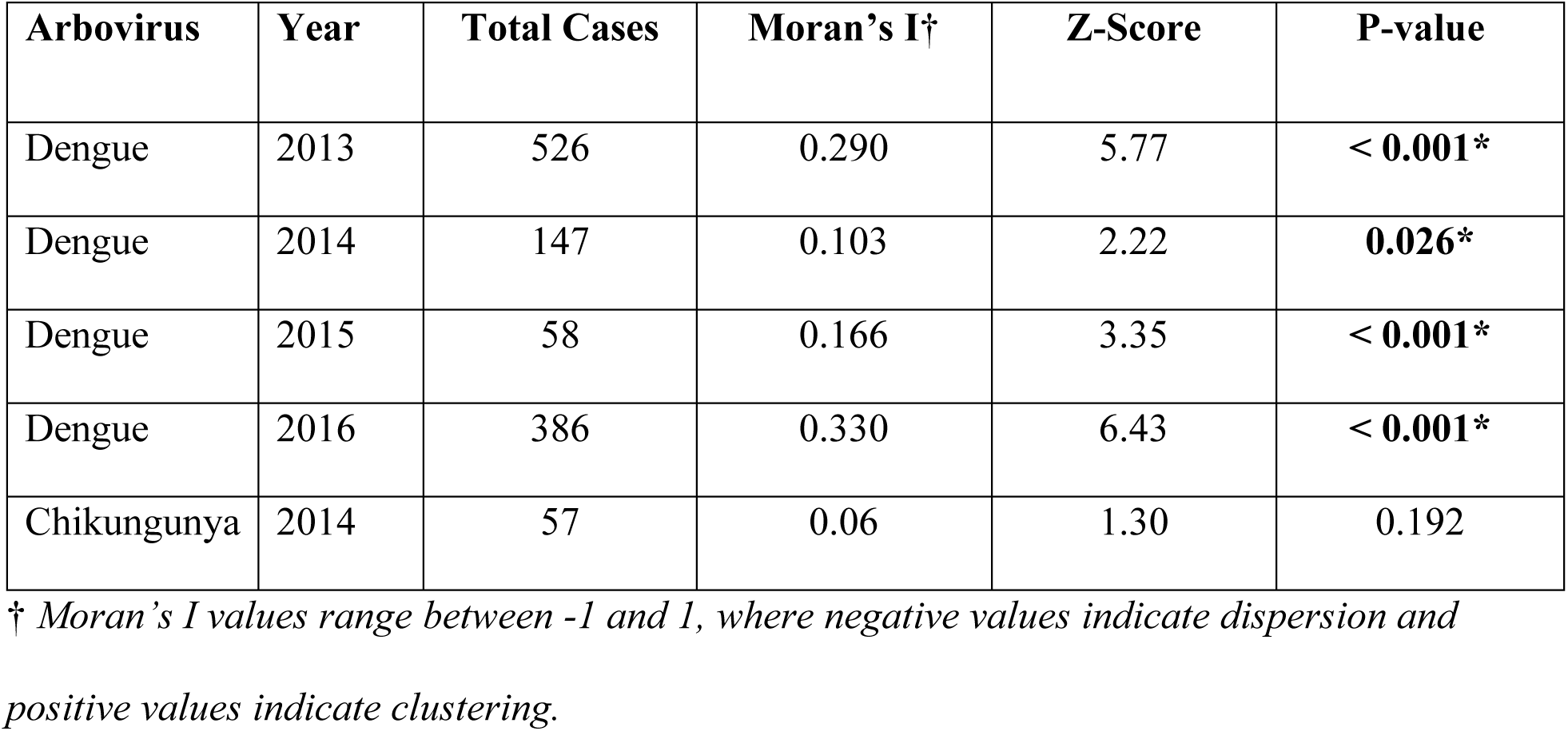
Global Moran’s I values for dengue and chikungunya EBS disease rates, aggregated to health district, in each year in the study.

| Arbovirus | Year | Total Cases | Moran's I<sup>†</sup> | Z-Score | P-value |
| --- | --- | --- | --- | --- | --- |
| Dengue | 2013 | 526 | 0.290 | 5.77 | < 0.001* |
| Dengue | 2014 | 147 | 0.103 | 2.22 | 0.026* |
| Dengue | 2015 | 58 | 0.166 | 3.35 | < 0.001* |
| Dengue | 2016 | 386 | 0.330 | 6.43 | < 0.001* |
| Chikungunya | 2014 | 57 | 0.06 | 1.30 | 0.192 |

<sup>†</sup> Moran's I values range between -1 and 1, where negative values indicate dispersion and positive values indicate clustering.

Local Moran’s I revealed shifting locations of dengue hotspots and coldspots at the health district level between years in both case totals (Fig. 2) and EB smoothed disease rates (Fig. 3). Localized spatial autocorrelation in dengue case counts was found during large outbreak years, while significant patterns of clustering in EB smoothed rates of dengue were found in every year.

**Fig. 2.**
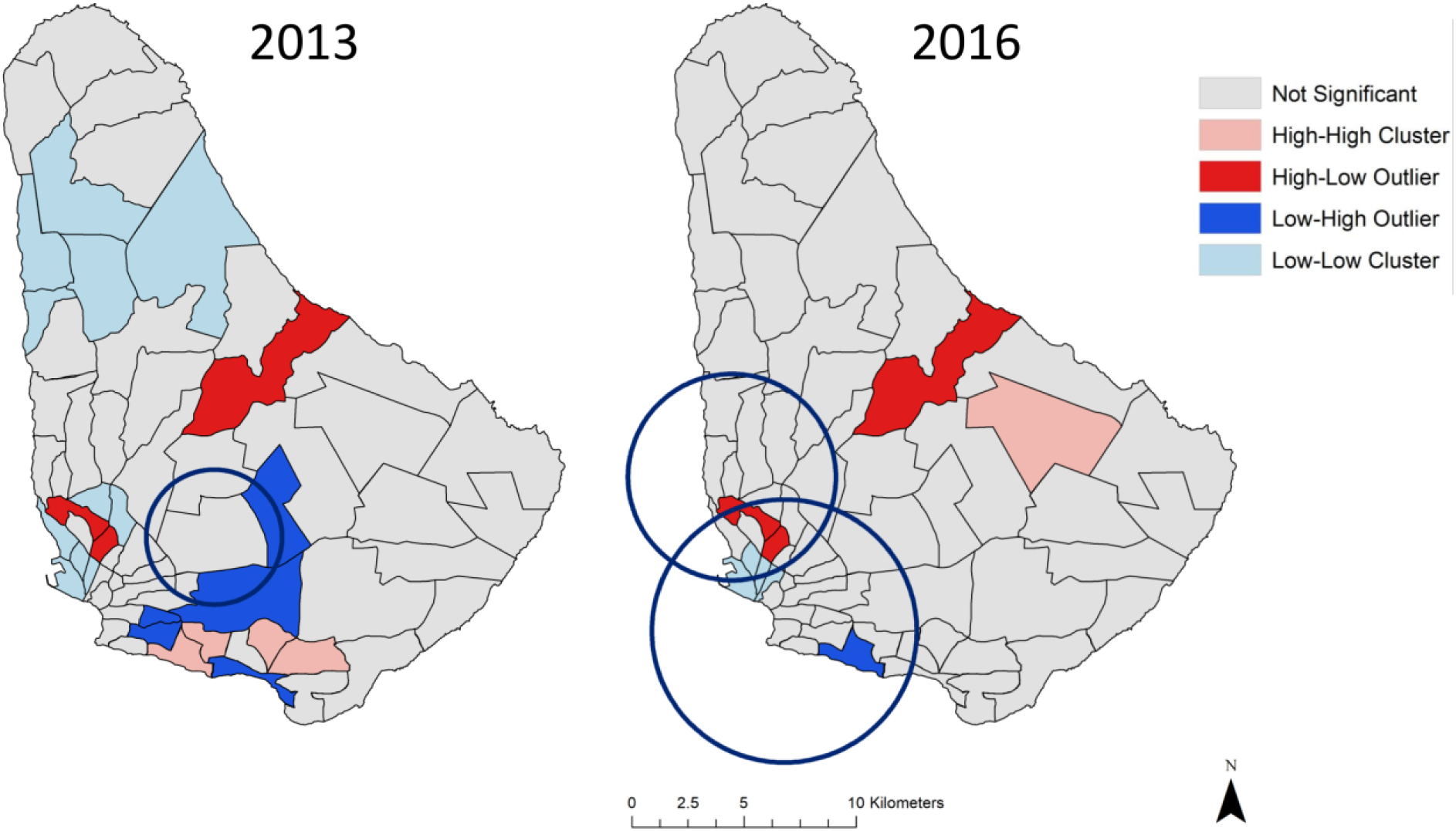
Patterns of clustering (red) and dispersion (blue) of dengue case totals were found at the level of health district in Barbados in 2013 and 2016, as determined by Local Moran’s I. Spatiotemporal coldspots (blue circles), found via the space-time spatial scan statistic, were found in both years. This figure was produced in ArcMap 10.4 (ESRI, Redlands, CA).

**Fig. 3.**
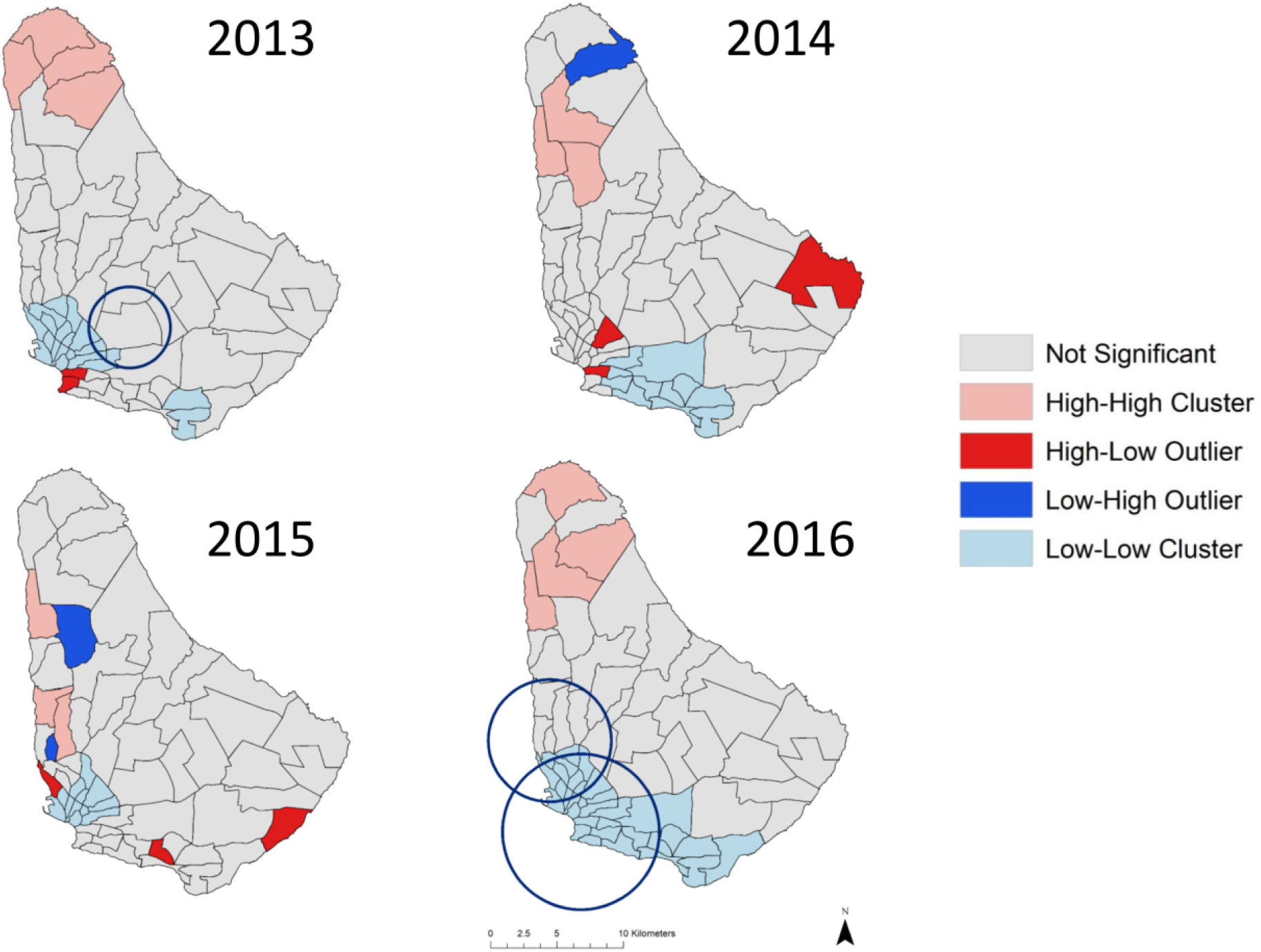
Patterns of clustering (red) and dispersion (blue) of EB smoothed dengue rates were found at the level of health district in Barbados for all years of the study, as determined by Local Moran’s I. Spatiotemporal coldspots (blue circles), found via the space-time spatial scan statistic, were found in 2013 and 2016 for dengue. This figure was produced in ArcMap 10.4 (ESRI, Redlands, CA).

The locations of hot and cold spots differed for case counts versus rates (Figs. 2 & 3). In years where both cases and rates had significant spatial autocorrelation (i.e. the 2013 and 2016 outbreaks), the highest disease rates were clustered in health districts in the north of the island, while dengue case counts had hotspots and clustered outliers (i.e. health districts with a high number of cases relative to neighboring districts with low counts) in central and southern health districts. Statistically significant spatial autocorrelation was only detected in smoothed rates in years of lower dengue burden (i.e. 2014 and 2015). During low burden years, hotspots were generally identified in northern health districts, while coldspots were detected in southern health districts (Fig. 3).

In years with large outbreaks of dengue, health districts in the southernmost Randall Philip PAC area were identified as hotspots of cases in 2013 (n=4), and in the centrally located St. Philip PAC area in 2016 (n=1) (Fig. 2). Coldspots for dengue cases were detected in health districts located in the Maurice Byer (n=5), Branford Taitt (n=4), and Eunice Gibson (n=2) PAC areas in 2013. In 2016, only 3 districts, in the Branford Taitt (n=2) and Winston Scott (n=1) administrative regions, were significant coldspots of cases (Fig. 2). Three health districts, located in Branford Taitt, Eunice Gibson, and David Thompson catchment areas, were found to be clustered outliers in both 2013 and 2016 (Fig. 2). When performing LISA analyses on smoothed dengue rates, the northern Maurice Byer PAC area contained all health districts that were hotspots of disease rates in 2013 (n=4) and 2016 (n=4) (Fig. 3). Significantly low rates of dengue were consistently found in southern health districts throughout the study period (Fig. 3).

Analysis of monthly case data in Barbados via spatial scan statistics did not identify statistically significant hotspots for either dengue or chikungunya. However, spatiotemporal coldspots of dengue cases were found in years with high case counts, indicating the duration and location of low disease activity during outbreak years (Table 3). In 2013, a coldspot spanning nine health districts across three PAC areas persisted from January to March (Table 3, Fig. 2). A large coldspot was also identified in 2016 from August-October, comprised of 29 health districts across five PAC areas, with a smaller, overlapping coldspot found in June to September of the same year.

**Table 3.** Statistically significant coldspots in monthly disease counts, calculated using the space-time permutation spatial scan statistic in SaTScan.

| Arbovirus | Year | PAC Areas | Health Districts | Duration (mm/yyyy) | Radius (km) | p-value |
| --- | --- | --- | --- | --- | --- | --- |
| Dengue | 2013 | SP, WS, EG | 9 | 01/2013 – 03/2013 | 3.03 | 0.034 |
| Dengue | 2016 | WS, RP, SP, BT, | 29 | 08/2016 – 10/2016 | 5.90 | < 0.001 |
| Dengue | 2016 | BT, EG, WS | 17 | 06/2016 – 09/2016 | 4.63 | 0.055* |

SP=St. Philip; WS=Winston Scott; EG=Eunice Gibson; RP=Randall Philip; BT=Branford Taitt

## Discussion

In this study, we found that cases of dengue fever in Barbados detected via surveillance in 2013–2016 exhibit both spatial and temporal structure. Dengue cases showed significant clustering in the central and southwestern health districts only in years with elevated case counts. In constrast, smoothed rates of population-derived incidence revealed clustering in all years for dengue, with many hotspots found in northern health districts. The identification of spatial dependence in disease cases is highly relevant for public health professionals working to suppress arbovirus transmission in Barbados, where there is a call to allocate public health resources efficiently.

Spatial discrepancies in data inputs (i.e. case numbers vs. population-derived rates) were driven in part by the small and spatially heterogeneous population density of Barbados. Consequently, any analyses performed on this system are susecptible to the “small numbers problem,” where estimates of commonly reported epidemiological metrics such as disease prevalence and incidence rates can dramatically fluctuate as an artefact of either low density of underlying populations, or relatively low case detection in high density populations ^26^. Procedures to reduce variance in rates, such as EB smoothing, are recommended to reduce the effect of unstable rates in disease mapping and tests for spatial autocorrelation ^29^. However, broad geospatial prescriptive remedies for the small numbers problem may unintentionally subvert public health agency management priorities, particularly in small island systems with extreme spatial population heterogeneity. Even after smoothing, we observed consistent hotspots of disease activity in northern health districts, where population densities are very low. Health districts with significantly high disease rates in low populations may not represent pragmatic management targets, especially in years where resources are limited or outbreaks are focused in urban centers. Although statistically sound, practical application of such analyses should be tempered by the expectations and priorities of public health agencies. In this context, raw case counts may give us a better understanding of operational disease burden on Barbados despite the problems typically associated with disregarding underlying population in morbidity metrics, where we would expect to detect more cases in densely populated areas regardless of true risk.

The differences observed in the spatial distribution of cases versus rates have critical implications with regards to intervention strategies and management goals. Although we accounted for inflated variance in rates by performing EB smoothing, hotspots in northern districts still reflect lower absolute case loads than found in densely populated areas in the south, especially in the vicinity of Bridgetown, the capital city. It is therefore imperative that management objectives are clearly specified before using spatial analyses on health surveillance data for planning purposes. Prioritization of goals is particularly important in a small island with high heterogeneity in population density, where making management decisions based on unstable rates could drive misallocation of resources. When responding to endemic transmission or emerging pathogens, like chikungunya, targeting areas with the highest transmission rates (i.e. high numbers of cases relative to the underlying population) should be prioritized to prevent further spread. Conversely, when considering large outbreaks of endemic diseases, like dengue, the management focus may be instead on reducing the total number of infections, regardless of population density, to mitigate hyperendemic peak years and reduce costs associated with the delivery of health services. In Barbados, these fundamental management distinctions may be subtle, but as our analyses demonstrate, can require vastly different spatial representations of disease clustering in the study area. This would directly translate to choices of where to allocate resources in particular health districts.

While we observed shifts in the clustering and dispersion of disease activity in Barbados between years, there were nevertheless consistencies in the location of health districts with clustered dengue cases or rates, especially in outbreak years. In particular, health districts identified as high clustering outliers during peak years were identical in 2013 and 2016, suggesting that some areas may have an underlying susceptibility to localized outbreaks when transmission is high (Fig. 2). Although the analyses presented here represent a reactive management approach, in which there is lagged decision-making in response to previously reported case data, spatial methodologies can also be incorporated into proactive strategies as part of an early warning system framework. Predictive climatological models of dengue risk have been developed for Barbados, enabling the anticipation of large outbreak events driven by environmental factors ^12^. Although useful in terms of triggering agency response ahead of major island wide outbreak events, current probabilistic forecast models do not provide information on ***where*** to intervene. Here, our spatial analyses point to consistent areas of transmission peaks, providing complementary analyses to predictive climate modeling efforts, which can be incorporated into MoH decision making, targeting discrete locations for mosquito control ahead of anticipated outbreak events.

We did not identify significant spatiotemporal clustering of dengue or chikungunya with spatial scan statistics within the study period, although coldspots of low dengue activity were found in years with higher case totals. Previous studies have indicated that spatial scan statistics are often more sensitive to the detection of hotspots, particularly when relative risk is low, compared to other exploratory methods of spatial analysis ^30,31^. Spatial scan statistics have also been used to successfully identify hotspots of mosquito-borne diseases at fine temporal resolutions in systems where diseases are endemic ^32,33^. Our inability to detect disease clustering at higher temporal resolutions, even in years with high case counts, perhaps points to a lack of within season localized clustering. The coldspots detected in 2016 for dengue coincide with the rainy season in Barbados, when we would expect to see increased transmission (Table 3). Coldspots arise as a result of spatial uniformity in risk outside these areas of unexpectedly low transmission. Our ability to detect spatial clustering at subseason scales may alternatively be hindered by human movements, reflecting the difficulty of performing local disease surveillance in transient populations (e.g. commuters and international travelers) ^34^. Although the inability to detect monthly clustering of arbovirus cases may limit the utility of spatial scan statistics to direct mosquito control activities at fine temporal scales in Barbados, our identification of coldspots during active transmission seasons warrants investigation and future research into potential drivers.

Aggregated surveillance data are routinely collected in a public health context, but are not free from limitations. The MoH verifies dengue and chikungunya cases in the laboratory, but underreporting of cases is a commonly encountered issue with passive surveillance data in Latin America and the Caribbean, resulting in an underestimation of true disease risk in some areas ^35^. The availability of current population data for calculating disease rates is also a limitation of this work. Although the projected population growth for Barbados is quite low, the most recent census data were collected in 2010, nearly a decade ago ^24^. In a spatial analysis context, aggregation of cases to health districts prevents us from drawing conclusions at finer scales. Although this limits our ability to inform household-level interventions within disease clusters, identifying health districts with high level of disease transmission is nevertheless relevant to the operational scale of health services delivered by the MoH in Barbados. Despite these limitations, the data used in this study represent the most accurate, and up-to-date estimates of population and disease risk in Barbados.

### Public Health Implications

These initial results serve as the foundation for incorporating spatial analyses into the existing arbovirus surveillance network in Barbados. Moving forward, these methodologies provide us not only with a means of guiding ministry responses to outbreaks of mosquito-borne diseases, but also the impetus for future geospatial analytical health studies in Barbados. Exploratory spatial analyses allow us to test hypotheses related to dominant social-ecological drivers of spatial clustering in health districts. Understanding the human characteristics that underlie observed spatial patterns can contribute to the development of better intervention methods.

## Data Availability

The data that support the findings of this study are available from the MoH, Barbados, but restrictions apply to the availability of these data, which were used under license for the current study, and so are not publicly available. Data are however available from the authors upon reasonable request and with permission of the MoH, Barbados.

## Declarations

### Authors’ Contributions

CAL, SJR, and AMS conceived of the study. CAL, AMS, MR, AQJH, RM, CJM, LR, MGH, ART, DH, and SK compiled the data used in analyses. CAL conducted analyses. CAL, SJR, and AMS drafted the manuscript. CAL, AMS, RL, RM, CJM, LR, MGH, ART, MJB, and SJR assisted with interpretation of the data and provided feedback for this manuscript. All authors read and approved the final manuscript.

## Literature Cited

1. Knudsen AB. Aedes aegypti and dengue in the Caribbean. AMCA Mosq News. 1983;43(4):269–275.

2. Brathwaite Dick O, San Martin JL, Montoya RH, del Diego J, Zambrano B, Dayan GH. The History of Dengue Outbreaks in the Americas. Am J Trop Med Hyg. 2012;87(4):584–593. doi:10.4269/ajtmh.2012.11-0770

3. Ryan SJ, Lippi CA, Carlson CJ, et al. Zika Virus Outbreak, Barbados, 2015–2016. Am J Trop Med Hyg. 2018;98(6):1857–1859. doi:10.4269/ajtmh.17-0978

4. Gittens-St Hilaire M, Clarke-Greenidge N. An analysis of the subtypes of dengue fever infections in Barbados 2003-2007 by reverse transcriptase polymerase chain reaction. Virol J. 2008;5(1):152. doi:10.1186/1743-422X-5-152

5. Gubler DJ. The Economic Burden of Dengue. Am J Trop Med Hyg. 2012;86(5):743–744. doi:10.4269/ajtmh.2012.12-0157

6. Halasa YA, Zambrano B, Shepard DS, Dayan GH, Coudeville L. Economic Impact of Dengue Illness in the Americas. Am J Trop Med Hyg. 2011;84(2):200–207. doi:10.4269/ajtmh.2011.10-0503

7. Shepard DS, Undurraga EA, Halasa YA, Stanaway JD. The global economic burden of dengue: a systematic analysis. Lancet Infect Dis. 2016;16(8):935–941. doi:10.1016/S1473-3099(16)00146-8

8. WTTC. Travel and Tourism Economic Impact 2018 Barbados. 2018.

9. WTTC. Travel and Tourism Economic Impact 2017 Barbados. 2017.

10. Shirtcliffe P, Cameron E, Nicholson KG, Wiselka MJ. Don’t forget dengue! Clinical features of dengue fever in returning travellers. J R Coll Physicians Lond. 1998;32(3):235–237.

11. Trotman A, Mahon R, Shumake-Guillemot J, Lowe R, Stewart-Ibarra AM. Strenghthening climate services for the health sector in the Caribbean. World Meteorol Organ. 2018;67(2).

12. Lowe R, Gasparrini A, Van Meerbeeck CJ, et al. Nonlinear and delayed impacts of climate on dengue risk in Barbados: A modelling study. Thomson M, ed. PLOS Med. 2018;15(7):e1002613. doi:10.1371/journal.pmed.1002613

13. Stewart-Ibarra AM, Muñoz Ángel G, Ryan SJ, et al. Spatiotemporal clustering, climate periodicity, and social-ecological risk factors for dengue during an outbreak in Machala, Ecuador, in 2010. BMC Infect Dis. 2014;14(1). doi:10.1186/s12879-014-0610-4

14. Lippi CA, Stewart-Ibarra AM, Muñoz ÁG, et al. The Social and Spatial Ecology of Dengue Presence and Burden during an Outbreak in Guayaquil, Ecuador, 2012. Int J Environ Res Public Health. 2018;15(4). doi:10.3390/ijerph15040827

15. Pepin KM, Marques-Toledo C, Scherer L, Morais MM, Ellis B, Eiras AE. Cost-effectiveness of Novel System of Mosquito Surveillance and Control, Brazil. Emerg Infect Dis. 2013;19(4):542–550. doi:10.3201/eid1904.120117

16. Paaijmans KP, Thomas MB. The influence of mosquito resting behaviour and associated microclimate for malaria risk. Malar J. 2011;10(1):183. doi:10.1186/1475-2875-10-183

17. Nathan MB. Critical review of Aedes aegypti control programs in the Caribbean and selected neighborhing countries. J Am Mosq Control Assoc. 1993;9(1-7).

18. McLafferty SL. GIS and Health Care. Annu Rev Public Health. 2003;24(1):25–42. doi:10.1146/annurev.publhealth.24.012902.141012

19. Oliveira MA de, Ribeiro H, Castillo-Salgado C, Oliveira MA de, Ribeiro H, Castillo-Salgado C. Geospatial analysis applied to epidemiological studies of dengue: a systematic review. Rev Bras Epidemiol. 2013;16(4):907–917. doi:10.1590/S1415-790X2013000400011

20. Castillo KC, Körbl B, Stewart A, Gonzalez JF, Ponce F. Application of spatial analysis to the examination of dengue fever in Guayaquil, Ecuador. Procedia Environ Sci. 2011;7:188–193. doi:10.1016/j.proenv.2011.07.033

21. Sugumaran R, Larson SR, DeGroote JP. Spatio-temporal cluster analysis of county-based human West Nile virus incidence in the continental United States. Int J Health Geogr. 2009;8(1):43. doi:10.1186/1476-072X-8-43

22. Kulldorff M. A spatial scan statistic. Commun Stat – Theory Methods. 1997;26. doi:10.1080/03610929708831995

23. Kulldorff M, SaTScan™ v7.0.3: Software for the spatial and space-time scan statistics. Information Management Services, Inc.

24. BSS. Barbados Population and Housing Censuss. 2010. http://www.barstats.gov.bb/census/.

25. WMO Caribbean Regional Climate Centre. http://rcc.cimh.edu.bb/. Published 2017. Accessed December 29, 2017.

26. Kennedy S. The small number problem and the accuracy of spatial databases. In: The Accuracy Of Spatial Databases. CRC Press; 1989:308. doi:10.1201/b12612

27. Anselin L. Local indicators of spatial association—LISA. Geogr Anal. 1995;27(2):93–115.

28. Kulldorff M, Heffernan R, Hartman J, Assunção R, Mostashari F. A Space–Time Permutation Scan Statistic for Disease Outbreak Detection. Blower SM, ed. PLoS Med. 2005;2(3):e59. doi:10.1371/journal.pmed.0020059

29. Olsen SF, Martuzzi M, Elliott P. Cluster analysis and disease mapping--why, when, and how? A step by step guide. BMJ. 1996;313(7061):863–866. doi:10.1136/bmj.313.7061.863

30. Barro AS, Kracalik IT, Malania L, et al. Identifying hotspots of human anthrax transmission using three local clustering techniques. Appl Geogr. 2015;60:29–36. doi:10.1016/j.apgeog.2015.02.014

31. Aamodt G, Samuelsen SO, Skrondal A. A simulation study of three methods for detecting disease clusters. Int J Health Geogr. 2006;5:15. doi:10.1186/1476-072X-5-15

32. Desjardins MR, Whiteman A, Casas I, Delmelle E. Space-time clusters and co-occurrence of chikungunya and dengue fever in Colombia from 2015 to 2016. Acta Trop. 2018;185:77–85. doi:10.1016/j.actatropica.2018.04.023

33. Coleman M, Coleman M, Mabuza AM, Kok G, Coetzee M, Durrheim DN. Using the SaTScan method to detect local malaria clusters for guiding malaria control programmes. Malar J. 2009;8(1):68. doi:10.1186/1475-2875-8-68

34. Walker AT, LaRocque RC, Sotir MJ. Travel Epidemiology. In: CDC Yellow Book. Centers for Disease Control and Prevention; 2019:720.

35. Torres JR, Orduna TA, Piña-Pozas M, Vázquez-Vega D, Sarti E. Epidemiological Characteristics of Dengue Disease in Latin America and in the Caribbean: A Systematic Review of the Literature. J Trop Med. 2017;2017:8045435. doi:10.1155/2017/8045435

